# A Protocol for a Scoping Study of Economic and Data System Considerations for Climate Change and Pandemic Preparedness in Africa

**DOI:** 10.1101/2024.02.23.24303199

**Authors:** Ariel Brunn, Francis Ruiz, Jane Falconer, Ellie Delight, Jessica Gerard, Yang Lui, Bernard Bett, Bubacarr Bah, Benjamin Uzochukwu, Kris Murray

## Abstract

**Background:** Africa’s changing climate heightens risk of disease outbreaks with impacts on vulnerable demographics already encumbered by health and socioeconomic burdens. Health preparedness policies to address disease outbreaks rely on integrated information systems and value-for-cost analysis to facilitate sound decision-making during a public health emergency. This rapid scoping review protocol describes the *a priori* objectives and methods to conduct a synthesis of the evidence on economic evaluation and data system capacity at the intersection of pandemic preparedness and climate change.

**Methods:** A search of six bibliographic databases will be conducted by a library information professional spanning the period 2010 – 2023, focused on literature published about pandemic preparedness in Africa in the context of climate change. Studies will be screened in a three-stage process by independent reviewers using Covidence software, with a proportion of excluded articles crosschecked by a reviewer not involved in screening. All articles included in the final study set will need to have a positive response to at least three out of four *a priori* screening questions. Data extraction will follow established frameworks for pandemic preparedness, a list of 14 target climate-sensitive infectious diseases with pandemic potential declared as Public Health Emergencies of International Concern and listed on the WHO’s R&D Blueprint Pathogens, and economic evaluation or data systems domains. Evidence synthesis will include article bibliometric analysis as well as thematic topic categorisation. Gap analysis will be conducted through topic mapping.

**Discussion:** This protocol lists the methods and analysis that will be followed to survey the literature on the linkages between climate change and pandemic preparedness relating to economic evaluation and data systems structures and needs.

## Background

Climate change exacerbates exposures and intensifies vulnerabilities for populations worldwide encumbered by health and socioeconomic burdens. In Africa, low-and-middle-income countries (LMICs) contribute nominally to greenhouse gas (GHG) emissions but are projected to face outsized impacts of climate change which threaten population health. An absence of effective and joined-up institutional cooperation and strong governance mechanisms limit opportunities for transdisciplinary and data-driven responses to complex health emergencies such as epidemic and pandemic outbreaks.

In conjunction with shifting land-use patterns, climate change both heightens risks for climate-sensitive infectious disease (CSID) emergence and spread and can lead to the re-emergence of previously controlled infectious diseases. Alterations in both habitat suitability and geographic range of wildlife species potentiate new pathogen emergence by driving cross-species pathogen exchange at high-risk interfaces (Carlson et al., 2022; Gibb et al., 2020) or through introduction of non-endemic diseases into new geographic areas and new transmission pathways (Ranjan et al., 2023; Thomassen et al., 2013). Favourable environmental conditions might also allow recrudescence of particular diseases such as sporulating or saprophytic pathogens, or facilitate re-introduction of disease vectors previously excluded by programs of countermeasures (Cohen et al., 2012; Walsh et al., 2018). Notably, all eight of the viral diseases ever declared by the World Health Organisation (WHO) as Public Health Emergencies of International Concern (PHEIC) and eight of nine^1^ diseases prioritised by the Research & Development (R&D) Blueprint for pathogens with epidemic potential (WHO, 2022) are categorised as climate-sensitive, according to a recent review (Mora et al., 2022). Socially marginalised demographic groups are most at risk from the unequal impacts of climate change, for instance flooding events may impede patient access to healthcare services causing delays to diagnosis or treatment (Zuurmond et al., 2016) with potential to raise the risk of uncontrolled disease spread and pandemic emergence.

While prevention and detection of emergence of CSIDs presents significant challenges, the spiralling costs of response and control measures associated with pandemic outbreaks due to long-lasting social, public health, and economic downturns, as seen in the 2014 Ebola outbreak in West Africa (Huber et al., 2018) and more recently with COVID-19, leave little doubt as to the necessity for established pandemic preparedness procedures. These recent outbreaks demonstrate the importance of agile and rapid decision-making by policy actors, reinforced by strong health governance and supported by integrated data systems and priority-setting processes based on value-for-cost assessments. Developing evidence-informed and data-driven policy responses builds confidence and public support for deployment of public funds during a public health crisis.

Economic considerations with respect to climate change and pandemic preparedness can be viewed through the lens of resource allocation choices which include exploring the cost-effectiveness of outbreak interventions and prevention. In this instance, economic evaluation (such as cost-effectiveness/cost-utility analysis, or cost-benefit analysis) is normally incorporated as part of priority-setting processes, which are ideally institutionalised and include approaches such as Health Technology Assessment (HTA) that consider a body of evidence and consider multi-stakeholder perspectives. Economic evaluation and priority-setting for CSIDs can be framed through an adapted framework outlining four stages of pandemic preparedness, namely, Prevention (Stage 1): pre-epidemic preparedness; detection (Stage 2): identify, investigate, evaluate risk; Response (Stage 3): outbreak response & containment; Evaluation (Stage 4): Post-epidemic evaluation (WHO, 2014). Successful priority setting structures requires capacity to exist both on the supply side in terms of evidence generation (e.g. the availability of skilled researchers able to undertake a relevant economic evaluation), and also on the demand side, such as the presence of formalised systems or bodies to use this evidence for policy.

As illustrated in Figure 1, Stage 1 of the adapted pandemic preparedness framework describes the emergence and re-emergence of infectious diseases with pandemic potential, whose emergence, distribution, or virulence can be affected by climate change. Literature that provides evidence of priority-setting methods, data collection systems, or economic evaluation of methods for the prevention of emergence/re-emergence of CSIDs will be categorised as Stage 1 evidence. In contrast, Stage 2 describes the effect of detection measures in preventing CSID outbreaks after emergence has occurred, including early warning surveillance and cost-effectiveness evaluations thereof. Stage 3 encompasses evidence on priority-setting processes and economic evaluation of outbreak response countermeasures. Finally, literature categorised as Stage 4 evidence will include post-outbreak and resource allocation analysis, such as after-action reviews.

**Figure 1.**
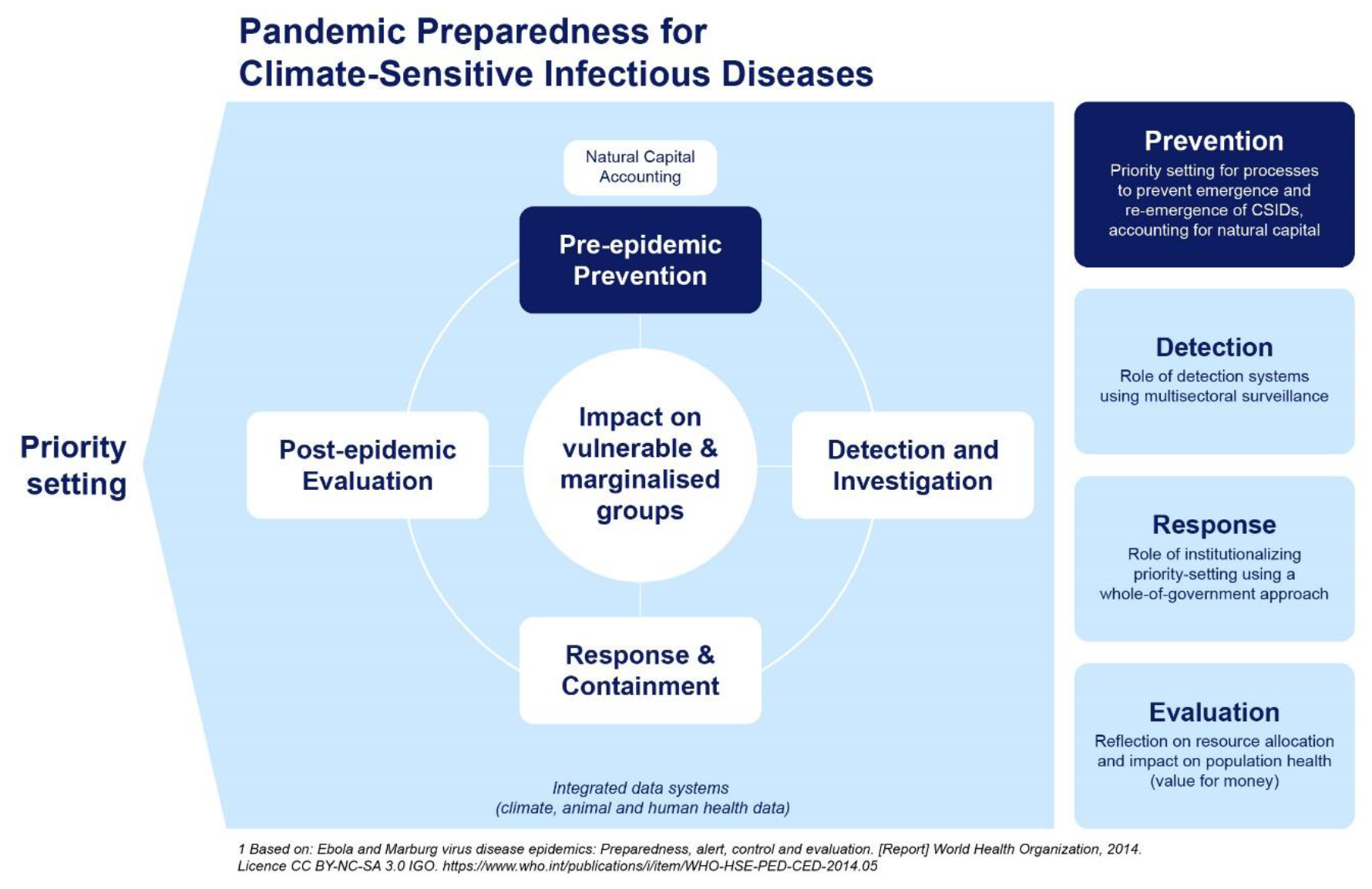
Pandemic Preparedness Framework for Climate Sensitive Infectious Disease in Africa. Adapted from: World Health Organisation. (2014). Ebola and Marburg virus disease epidemics: Preparedness, alert, control, and evaluation. https://www.who.int/publications-detail-redirect/WHO-HSE-PED-CED-2014.05

### Rationale

There is an urgent need to map evidence gaps, define future research priorities, and strengthen stakeholder networks at the climate-health interface to support policy development for better health outcomes and improved health systems resilience for vulnerable populations. This requires multidisciplinary and whole-of-government approaches, drawing from the integrative fields of One Health and Planetary Health, with a focus on health equity to prioritise marginalised demographics. By improving understanding of the climate-health nexus, strategies to prevent, detect, and respond to disease outbreaks can be developed more effectively.

Scoping reviews offer a systematic method of mapping the state of knowledge to identify common themes and to uncover gaps in evidence. Rapid reviews condense these methods and are often used to accelerate the knowledge translation process when policy or strategy development is urgently needed (Haby et al., 2016). This rapid scoping review protocol thus outlines the process used to define key themes and identify evidence gaps on economic evaluation and data systems in relation to pandemic preparedness and to develop a research agenda that supports progress on CSID knowledge and policy development in Africa.

### Scoping Review Objectives

i. To map out key themes and entry points for future African-led research and to identify gaps in evidence for priority setting using economic evaluation and data systems for policy development;
ii. To use bibliometric analysis to assess institutional stakeholder networks and to evaluate trends in research development on key themes and outputs included in the review;
iii. To identify applications for a transdisciplinary and gender-intersectional approach for CSID preparedness and response.

### Research Questions

i. What are the key themes, in terms of economic evaluation (including cost-effectiveness) and priority setting, that describe how climate change is incorporated into pandemic preparedness and outbreak response planning?
ii. What data systems are currently in use or needed to support priority setting for integrated and multisectoral epidemic or pandemic planning for climate-sensitive infectious diseases in Africa?
iii. What subnational, national, and regional structures are in place to support joined-up data sharing for CSID outbreak detection and response within and between African countries and the international community?

### Protocol

Informed by the Preferred Reporting Items for Systematic Reviews and Meta-Analyses (PRISMA) for Protocols (Shamseer et al., 2015) we outline our methods *a priori* for this rapid scoping review. The protocol is registered online on the Open Science Framework repository (https://doi.org/10.17605/OSF.IO/VTGMC).

**Table 1.** Scoping Review Eligibility Criteria.

| Criteria | Inclusion | Exclusion |
| --- | --- | --- |
| **Publication Period** | 2010 - 2023 | Articles outside this period. |
| **Location** | Articles relevant to and published in or about countries or regions in Africa. | Articles outside of Africa, articles not geographically located, or articles on globally aggregated data. |
| **Language** | Articles in English. | Non-English language articles will be excluded. |
| **Study Type** | Original research, literature reviews, reports, policy briefs, conference abstracts, opinion pieces and editorials. | News articles and conference abstracts |
| **Pandemic Preparedness** | Articles that assess preparedness that can be contextualised within the WHO (2014) Pandemic Preparedness Framework. | NA |
| **Climate Change** | Articles that refer to infectious diseases impacted by climate change; this encompasses weather variables, hydrometeorological hazards and land-use change over short or long (decadal) time spans.<br>CSIDs of zoonotic origin that have developed a human-to-human transmission cycle are included even if the | Articles that consider climate change-related hazards such as extreme weather as a barrier to healthcare facility access will be excluded.<br>Articles that refer to climate in a cultural sense, i.e. political climate, will be excluded.<br>Articles about allergens, fungal diseases and AMR are excluded in this review, although it is acknowledged |
|  | subsequent transmission is no longer sensitive to climate covariates. CSIDs with pandemic potential are adapted from the R&D Blueprint Disease List (WHO, 2022) and WHO PHEIC diseases (Wilder-Smith & Osman, 2020) | that these disease processes are increasingly being linked to climate change. |
| **Health Economic Theme** | Articles that describe the use of health technology assessment (HTA), priority-setting, cost effectiveness or cost-benefit analysis and pandemic preparedness in the context of climate change. | Articles that do not provide analysis or discussion in the context of pandemic preparedness or climate change. |
| **Data Systems Theme** | Articles that describe, propose, or analyse collaborative transdisciplinary data sharing systems including meteorological or animal health data systems centred on enhancing infectious disease detection, surveillance, and control. | Articles that mention data or information systems not relevant to a health or health economics context. |
| **Outcomes** | Descriptions of frameworks, data system or health information system structures, case study outcomes, trial outcomes, best practices, evidence syntheses including meta-analyses results and cost-effectiveness findings. Specific outcomes extracted include: study population, location of research, interventions, timeframe and evaluation methods and results. | Global economic evaluations and global meta-analyses where African data is not extractable. |

### Search Strategy

The search strategy will be compiled by a library information professional with a focus on four search concepts, namely:

- Pandemic preparedness AND
- Climate change AND
- Economic evaluation (including health technology assessment and priority-setting) OR
- Data systems

A draft search, run on the OvidSP Medline database is included as an appendix.

### Information Sources

Relevant literature will be included through searches of six bibliographic databases: OvidSP Medline, OvidSP Embase, OvidSP Global Health, EBSCOhost Africa-Wide Information, OvidSP Econlit and Clarivate Analytics Web of Science Core Content. Search terms will first be tested in one database prior to implementation in the other five. Additional sources of literature from hand-searching relevant reference lists and from work previously conducted by co-authors will be included.

Grey literature will be incorporated through review of relevant reports, documents, and publications from the International Development Research Centre (IDRC) and by asking key stakeholders for recommended literature, particularly non-indexed reports, through informal interviews taking place in parallel to the scoping review.

### Data Management

De-duplicated citations of bibliographic database and grey literature searches will be uploaded into Covidence software (Veritas Health Innovation, 2024) for reference management and multi-reviewer screening. Eligibility criteria will be used to identify articles for inclusion as described in the Data Selection process. All reviewers will be trained on the use of screening software.

### Data Selection

Articles from the bibliographic database and grey literature search will be compiled and put through three-stage screening. An initial stage of title screening will be conducted to exclude articles that can be quickly disregarded at the title stage. The second stage of screening on the remaining articles will select eligible abstracts against a set of four screening questions. All abstracts must have a positive response to screening questions number 1 and 2 and at least one of either question 3 or 4 to be eligible for inclusion in the final full-text screening stage. Screening will be conducted by single independent reviewers and a proportion of excluded articles will be randomly selected for confirmation by one member of the review team. Abstracts in languages other than English will be documented but excluded; translations will not be sought for full-text articles not in English. Reasons for study exclusion will be documented at both the abstract and the full-text screening stage for future summary. Studies that are inconclusive on screening will be discussed by the scoping review team to reach consensus on eligibility.

### Screening Questions

1. *Is the article relevant to pandemic preparedness?*
2. *Is the article about a climate-sensitive infectious disease(s) that is included in the target list of diseases?*
3. *Does the article refer to economic evaluation or health economic studies to support priority setting including Health Technology Assessment?*
4. *Does the article discuss data systems relevant to identified domains for priority setting (see Table A1)*.

### Data Collection Process

Data collection will use a standard form in Covidence, which will be formatted as a table with data items listed in columns to be populated for each article. The form will be pre-tested against ten articles in duplicate prior to use.

### Data Items

Data collected from each article will include article meta-data and article themes. Article meta-data will include year of publication, publication source (institution and country), first author and first author’s institution, collaborating institutions and contributors, source of funding, conflicts of interest reported, type of publication. Topic data will also be extracted and thematically categorised based on identified frameworks of the review. All articles must fit into one of the identified stages of the Pandemic Preparedness Framework (WHO, 2014). CSIDs will be identified and listed to confirm which diseases with pandemic potential are most researched. Data systems will be further sub-categorised as to the type of health information they incorporate across human, animal, or climate data, accessibility and overall degree of multi-disciplinarity; gender disparities will be captured in the thematic assessment phase. A final mapping of challenges and constraints will be conducted by assessing topics with the least volume or least relevant literature, and by identifying repeated themes describing evidence gaps arising in the final study set.

### Risk of Bias

A risk of bias assessment will not be conducted in this rapid scoping review; therefore, the authors will not be able to qualify that the conclusions are not biased. However, the authors will extract and summarise study methodologies which may be useful as an indicator of potential bias of evidence. Additionally, exclusion of non-English articles may bias the output to English-speaking countries in Africa, however, is a necessary exclusion criterion given the rapid nature of the scoping review.

### Synthesis of Results

The scoping review process used to achieve the final study set will be illustrated using a flow chart template. The number of studies at each stage of screening will be included and reasons for exclusion will be provided at both the title and abstract and the full-text screening stage. Extracted data will be analysed through three methods: bibliometric analysis to characterise the set of included articles; topic mapping to identify the key themes relevant to the review and identify evidence gaps; and narrative synthesis to contextualise the findings of the review.

### Bibliometric analysis

Bibliometric analysis is a methodology from the field of information science that is useful for identifying trends in large volumes of literature. In this review, we will use bibliometric analysis on tabulated article meta-data to define research hubs in Africa through institutional network analysis, to understand temporal trends in publication intensity, and to provide a summary of funding and research funders active at the intersection of pandemic preparedness and climate change. Both temporal and geographical trends will be assessed. We will attempt to explore authorship patterns to identify key advocates and networks of research on the African continent. We will also assess institutional affiliations of first authorship and corresponding authors as a measure of academic capacity, retention and leadership in the field.

### Topic mapping

Thematic topic categorisation will precede gap analysis. Evidence from the literature will be classified by its relevance to the four stages of the adapted Pandemic Preparedness Framework; by the share of research on each CSID; and by which sub-theme of economic evaluation or data system the article provides evidence for. Sub-themes will be categorised and mapped temporally and geographically, data permitting. Gap analysis will be conducted based on differences between research intensity per sub-theme category, as well as identifying institutional or regional discrepancies in evidence availability.

### Narrative synthesis

Articles will undergo simple narrative synthesis to describe the main objectives and findings which will be tabulated for use as a data reference of included articles. Narrative synthesis will also include determining the proportion of articles that consider transdisciplinary and multisectoral approach to data and information systems (“One Health” approach) and the proportion of articles that consider gender-intersectional themes.

### Ethics and dissemination

This review will use publicly available reports and articles from bibliographic databases and online. As no primary or identifiable data will be used, ethical approval is not required.

This scoping review protocol and the findings of the review itself will be shared with stakeholders and decision-makers and made available as an open-access report. Dissemination is anticipated through publication online and in peer reviewed journals as well as at relevant meetings and workshops.

### Scoping Study Team

Ariel Brunn, Department of Population Health, London School of Hygiene and Tropical Medicine, Keppel Street, London, WC1E 7HT, UK

Yang Liu, Department of Population Health, London School of Hygiene and Tropical Medicine, Keppel St, London WC1E 7HT, UK

Bernard Bett, International Livestock Research Institute, CGIAR

Francis Ruiz, Department of Global Health & Development, Faculty of Public Health and Policy, London School of Hygiene and Tropical Medicine, Keppel St, London WC1E 7HT, UK

Benjamin Uzochukwu, Department of Community Medicine, University of Nigeria Nsukka, Enugu campus, Nigeria

Kris Murray, MRC Unit The Gambia at London School of Hygiene and Tropical Medicine, Atlantic Boulevard, Fajara, The Gambia

Jane Falconer, Library, Archive and Open Research Services, London School of Hygiene and Tropical Medicine, Keppel St, London WC1E 7HT, UK.

Ellie Delight, MRC Unit The Gambia at London School of Hygiene and Tropical Medicine, Atlantic Boulevard, Fajara, The Gambia

Jessica Gerard, Faculty of Infectious and Tropical Diseases, London School of Hygiene and Tropical Medicine

Bubacarr Bah, MRC Unit The Gambia at London School of Hygiene and Tropical Medicine, Atlantic Boulevard, Fajara, The Gambia

### Support

This work is supported by the International Development Research Centre (Project Reference: 29957). The protocol was developed independently from the International Development Research Centre.

## Data Availability

All data produced are available online via the respective bibliographic databases: OvidSP Medline, OvidSP Embase, OvidSP Global Health, EBSCOhost Africa-Wide Information, OvidSP Econlit and Clarivate Analytics Web of Science Core Content.

---

**Appendix: Table A1.**
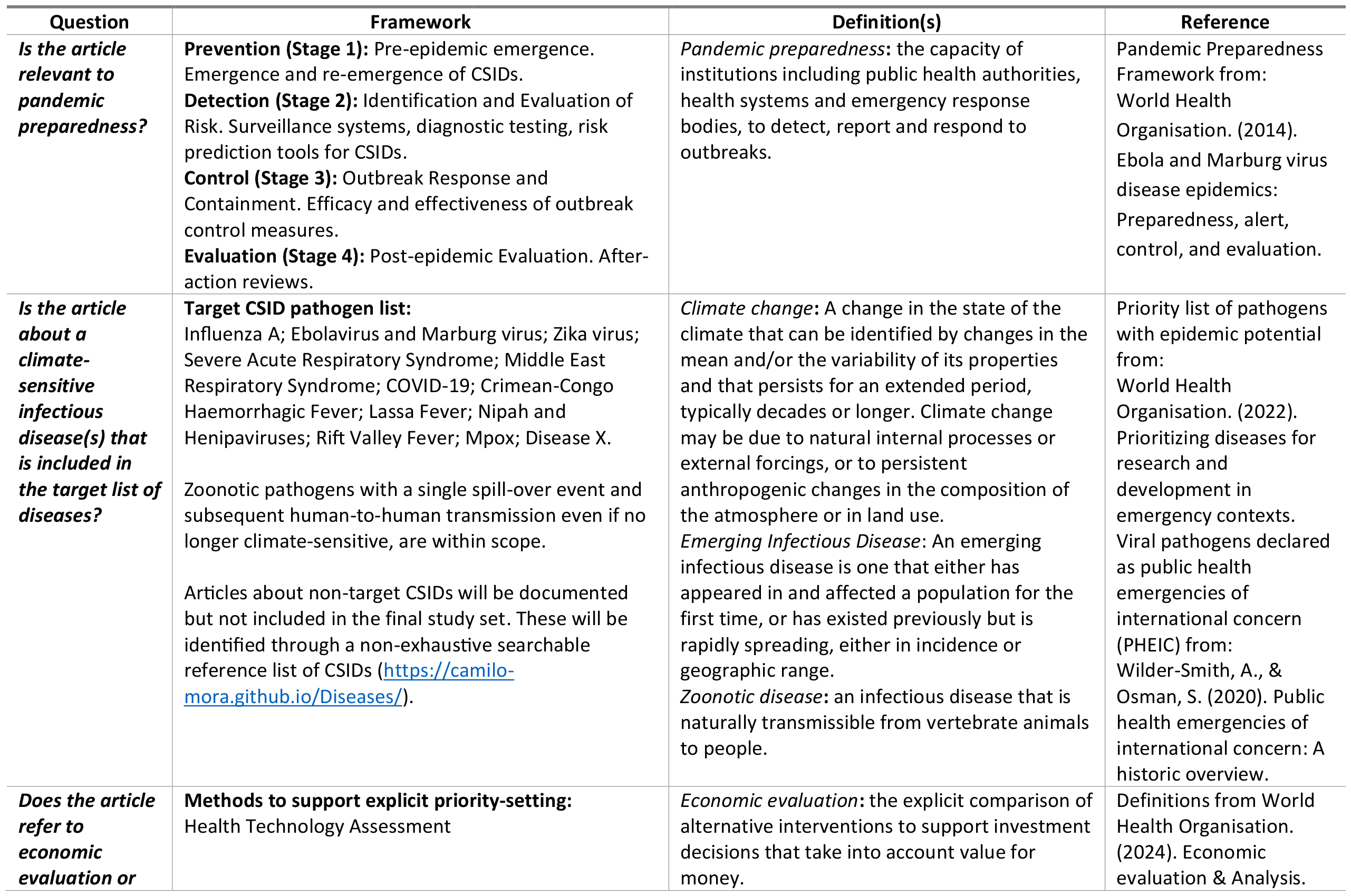
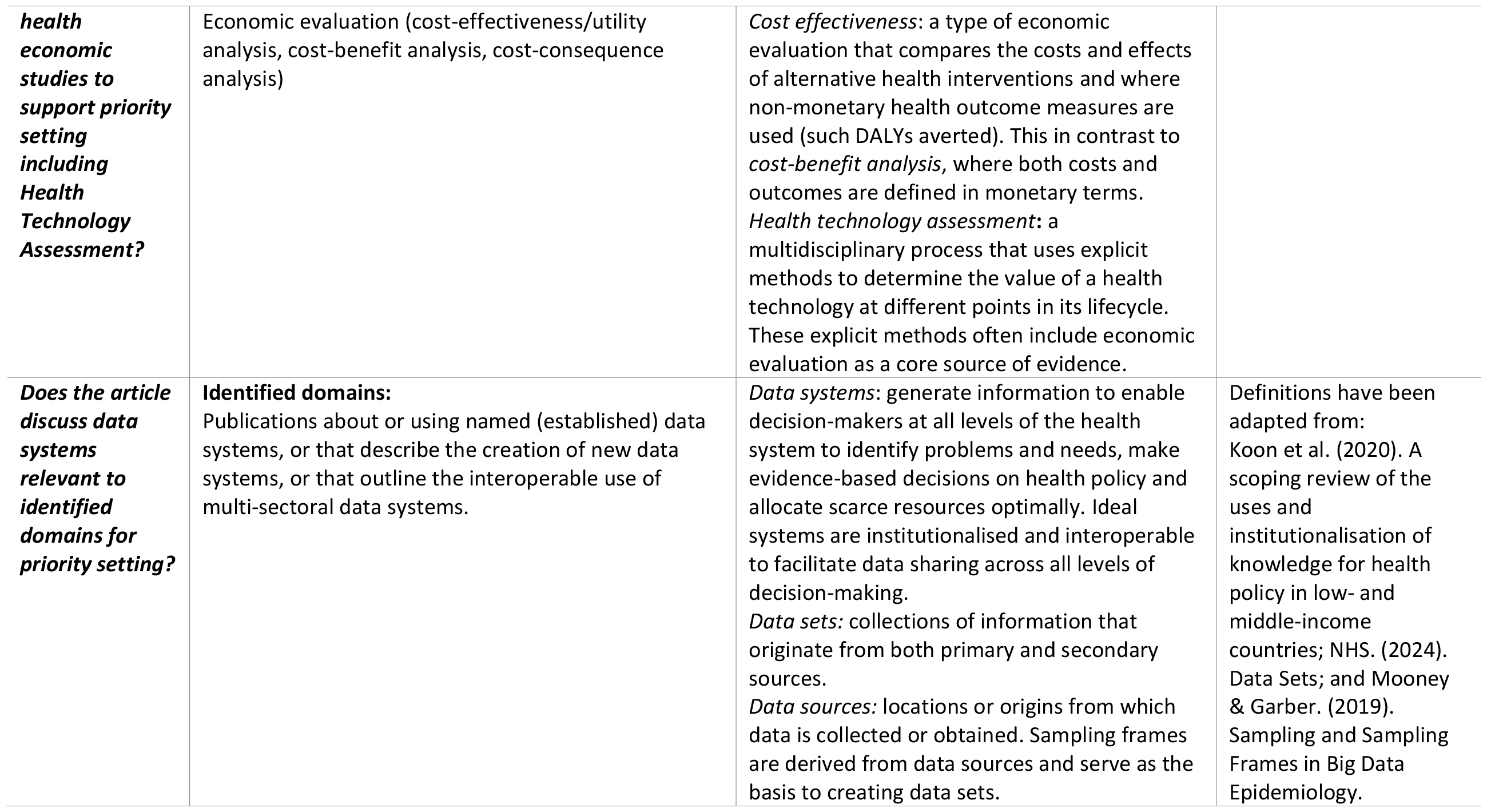
Screening Guidance.

### Appendix: OvidSP Medline draft search strategy

1. exp Human Activities/ and (ecosystem/ or exp ecology/ or environment/)
2. ((environment* or ecosystem* or ecolog*) adj3 (change* or variab* or disrupt* or hazard* or impact* or vulnerab* or function* or footprint* or function* or extreme or exposure or exposed or expose or sustainab*)).ti,ab.
3. ((environment* or global or planet* or ecolog*) adj1 (sustainab* or regenerat*)).ti,ab.
4. (climat* adj3 solution*).ti,ab.
5. exp Forests/
6. exp Tundra/
7. Wetlands/
8. exp Fresh Water/
9. Wilderness/
10. Rivers/
11. Parks, recreational/
12. (forest* not (random survival forest or forest plot*)).ti,ab.
13. (tree? not decision tree).ti,ab.
14. woodland?.ti,ab.
15. (grass not (grass-carp or grass-root?)).ti,ab.
16. (greenspace? or green-space?).ti,ab.
17. green-infrastructure.ti,ab.
18. park?.ti,ab.
19. tundra.ti,ab.
20. permafrost.ti,ab.
21. wetland?.ti,ab.
22. fresh-water.ti,ab.
23. (bluespace? or blue-space?).ti,ab.
24. lake?.ti,ab.
25. pond?.ti,ab.
26. (dam or dams or reservoir*).ti,ab.
27. river?.ti,ab.
28. wilderness.ti,ab.
29. desert?.ti,ab.
30. “the bush”.ti,ab.
31. coast*.ti,ab.
32. beach*.ti,ab.
33. Biodiversity/
34. Endangered species/
35. Introduced species/
36. Extinction, biological/
37. biodivers*.ti,ab.
38. biological divers*.ti,ab.
39. (extinct* adj2 (rate? or species)).ti,ab.
40. ((wildlife or species) adj2 (protect* or declin* or abundan* or rich*)).ti,ab.
41. protected area?.ti,ab.
42. biosphere.ti,ab.
43. exp Climate Change/ or Global Warming/
44. (exp Weather/ or exp Climate/) and exp Disasters/
45. ((climate or climactic) adj3 (change? or changing or variab* or disrupt* or hazard* or impact* or vulnerab* or function* or extreme or catastroph* or emergenc* or shift* or dynamic* or sensitiv* or sustainab* or anomal*)).ti,ab.
46. ((hydrometeorolo* or atmosphere* or hydrologic* or ocean* or weather) adj3 (change* or variab* or disrupt* or hazard* or extreme or exposure or exposed or expose or phenomen* or sustainab* or anomal*)).ti,ab.
47. extreme weather.ti,ab.
48. natural hazard?.ti,ab.
49. global tipping point?.ti,ab.
50. exp Temperature/
51. Ice Cover/
52. Permafrost/
53. ((global or planet* or world or climat* or earth or worldwide or world-wide or ambient air or ocean*) adj1 (heating or warming or temperature*)).ti,ab.
54. ((temperature? or cold or heat or hot or ambient air or relative humidity or thermal) adj4 (extreme or elevate* or irregular or intense or reduced or pattern* or anomal* or global)).ti,ab.
55. (ENSO or el nino-southern oscillation).ti,ab.
56. thermal threshold?.ti,ab.
57. thermal stratification.ti,ab.
58. ((arctic or antarctic or pack-ice or ice-sheet or glaci* or snow*) adj3 (shrink* or decrease* or melt*)).ti,ab.
59. (heatwave? or heat-wave?).ti,ab.
60. (coldwave? or cold-wave?).ti,ab.
61. (thermal environ* or heat-island*).ti,ab.
62. Floods/
63. Droughts/
64. ((sea-level or sea level) adj5 (rise? or rising or change? or changing)).ti,ab.
65. cloud cover.ti,ab.
66. ((rain* or precipitation) adj4 (extreme or elevate? or elevating or irregular or intense or reduced or pattern* or anomal*)).ti,ab.
67. flood*.ti,ab.
68. water current?.ti,ab.
69. water stress.ti,ab.
70. drought?.ti,ab.
71. Cyclonic Storms/
72. tropical cyclone?.ti,ab.
73. hurricane?.ti,ab.
74. storm*.ti,ab.
75. ((tide or tidal or sea or ocean*) adj2 (surge or surges)).ti,ab.
76. ((ocean* or sea or marine) adj2 (ph or acidification or heating or warming or change* or variab* or extreme or expand or expansion or sustainab*)).ti,ab.
77. windstorm?.ti,ab.
78. Landslides/
79. Wildfires/
80. landslide?.ti,ab.
81. (wildfire? or wild-fire?).ti,ab.
82. ((fire? or burnt or inferno or conflagration) adj2 (change* or variab* or extreme or expand or expansion or intense or elevate? or increas*or catastroph* or emergenc* or shift* or dynamic* or frequen* or more)).ti,ab.
83. Water Supply/
84. exp Groundwater/
85. (water adj3 footprint).ti,ab.
86. (global freshwater or global fresh-water).ti,ab.
87. global water cycle.ti,ab.
88. ((freshwater or water) adj1 (provision or provid* or supply or sustain*)).ti,ab.
89. (groundwater or ground-water).ti,ab.
90. aquifer*.ti,ab.
91. water table.ti,ab.
92. watershed.ti,ab.
93. (water adj4 (resource* or conserv* or security or availab* or scarcity or poverty or stress or overconsumpt* or over-consumpt* or save or saving or sustainab*)).ti,ab.
94. hydrological flow*.ti,ab.
95. soil erosion/
96. (land supply or land supplies).ti,ab.
97. “land use change?”.ti,ab.
98. (habitat adj2 (loss or fragment* or disturb* or alter or altered or destruct*)).ti,ab.
99. soil erosion.ti,ab.
100. soil quality.ti,ab.
101. land cover change*.ti,ab.
102. nutrient run-off.ti,ab.
103. deforestation.ti,ab.
104. land degradation.ti,ab.
105. “slash-and-burn”.ti,ab.
106. desertificat*.ti,ab.
107. Greenhouse Effect/
108. Greenhouse gases/
109. ozone depletion/
110. (greenhouse effect* or green-house effect*).ti,ab.
111. (greenhouse gas* or green-house gas*).ti,ab.
112. ((“co2” or carbon or methane or ozone or nitrous oxide) adj5 (elevat* or emission* or rise or increas* or decreas* or reduc* or neutral or offset* or sustainab*)).ti,ab.
113. ((global or planet* or world or earth or worldwide or world-wide) adj2 (“co2” or carbon or methane or ozone or nitrous oxide)).ti,ab.
114. (emission* adj5 (elevat* or rise or increas* or decreas* or reduc* or offset* or sustainab*)).ti,ab.
115. carbon footprint/
116. (carbon adj3 footprint).ti,ab.
117. (de-carbon or de-carboni* or decarbon or decarboni*).ti,ab.
118. exp water pollution/
119. (water adj2 (pollut* or contamina*)).ti,ab.
120. water quality.ti,ab.
121. exp waste products/
122. waste.ti,ab.
123. rubbish.ti,ab.
124. trash.ti,ab.
125. garbage.ti,ab.
126. fly-tipping.ti,ab.
127. dump*.ti,ab.
128. sewage.ti,ab.
129. (faeces or faecal or feces or fecal).ti,ab.
130. excrement.ti,ab.
131. slurry.ti,ab.
132. “Environmental Restoration and Remediation”/
133. cleanup?.ti,ab.
134. environmental remediation?.ti,ab.
135. environmental restoration.ti,ab.
136. pollution remediation?.ti,ab.
137. Sanitation/
138. sanitat*.ti,ab.
139. 139 or/1-138
140. disease outbreaks/
141. epidemics/
142. pandemics/
143. pandemic*.ti,ab.
144. epidemic*.ti,ab.
145. (public health emergenc* or PHEIC).ti,ab.
146. (disease adj1 spread*).ti,ab.
147. outbreak*.ti,ab.
148. (climat* adj3 (infect* or communicab* or diseas*)).ti,ab.
149. (geograph* adj3 diseas*).ti,ab.
150. Influenza A Virus, H1N1 Subtype/
151. 151 “h1n1”.ti,ab.
152. Hemorrhagic Fever, Ebola/ or Ebolavirus/
153. (haemorrhagic fever or hemorrhagic fever or ebola or ebolavirus).ti,ab.
154. Zika Virus/ or Zika Virus Infection/
155. zika.ti,ab.
156. Severe acute respiratory syndrome-related coronavirus/ or coronavirus infections/ or severe acute respiratory syndrome/
157. Middle East Respiratory Syndrome Coronavirus/
158. COVID-19/ or SARS-CoV-2/
159. (sars* or severe acute respiratory syndrome).ti,ab.
160. (mers* or middle east respiratory syndrome).ti,ab.
161. (covid-19 or (“2019” adj2 (coronavirus* or ncov)) or wuhan coronavirus).ti,ab.
162. Hemorrhagic Fever, Crimean/
163. Hemorrhagic Fever Virus, Crimean-Congo/
164. ((crimean or congo) adj2 (hemorrhagic or fever or virus)).ti,ab.
165. Lassa Fever/ or Lassa virus/
166. (lassa adj1 (fever or virus or infect*)).ti,ab.
167. exp Henipavirus/ or Henipavirus Infections/
168. (nipah or henipav*).ti,ab.
169. Rift Valley fever virus/ or Rift Valley Fever/
170. rift valley fever.ti,ab.
171. (disease x not x-linked).ti,ab.
172. or/140-171
173. exp Disaster planning/
174. exp Communicable Disease Control/
175. exp Technology Assessment, Biomedical/
176. exp Health Planning/
177. exp Regional Health Planning/
178. exp Costs/ and Cost Analysis/
179. exp Economics/
180. exp Evidence-Based Practice/
181. decision making/
182. clinical decision making/
183. health equity/
184. exp records/
185. exp algorithms/
186. data systems/
187. database management systems/
188. electronic data processing/
189. exp computer security/
190. exp data science/
191. exp data management/
192. exp data mining/
193. data compression/
194. exp information management/
195. (preparedness or response).ti,ab.
196. (prevent* adj1 control).ti,ab.
197. ((infection or disease* or measure*) adj1 control).ti,ab.
198. (health technology assessment or HTA).ti,ab.
199. (economic* or economy or economies).ti,ab.
200. (macroeconomic* or macro-economic*).ti,ab.
201. (priorit* adj1 (health* or set* or area*)).ti,ab.
202. (resourc* adj1 allocat*).ti,ab.
203. (resourc* adj3 distrib*).ti,ab.
204. logistic*.ti,ab.
205. surge capacit*.ti,ab.
206. ((health* or resourc*) adj3 ration*).ti,ab.
207. cost-effectiv*.ti,ab.
208. (cost-benef* adj1 analys*).ti,ab.
209. (cost-utilit* adj1 analys*).ti,ab.
210. (cost or costs).ti,ab.
211. (financ* or fiscal*).ti,ab.
212. marginal analys*.ti,ab.
213. (benefit* adj2 cost*).ti,ab.
214. (evidenc* adj2 (assess* or apprais* or eval*)).ti,ab.
215. (evidenc* adj1 base*).ti,ab.
216. best-practic*.ti,ab.
217. (decision* adj2 mak*).ti,ab.
218. (health* adj1 equit*).ti,ab.
219. coordinat*.ti,ab.
220. data.ti,ab.
221. information*.ti,ab.
222. or/173-221
223. exp Africa/
224. africa.ti,ab.
225. african countr*.ti,ab.
226. AFRO.ti,ab.
227. Algeria.ti,ab.
228. (Djibouti or French Somaliland).ti,ab.
229. Egypt.ti,ab.
230. Morocco.ti,ab.
231. Tunisia.ti,ab.
232. Libya.ti,ab.
233. Angola.ti,ab.
234. (Cameroon or Kamerun or Cameroun).ti,ab.
235. (Cape Verde or Cabo Verde).ti,ab.
236. (Comoros or Glorioso Islands or Mayotte).ti,ab.
237. (Congo not ((Democratic Republic adj3 Congo) or congo red)).ti,ab.
238. (Cote d’Ivoire or Cote dIvoire or Ivory Coast).ti,ab.
239. (eSwatini or Swaziland).ti,ab.
240. (Ghana or Gold Coast).ti,ab.
241. (Kenya or East Africa Protectorate).ti,ab.
242. (Lesotho or Basutoland).ti,ab.
243. Mauritania.ti,ab.
244. Nigeria.ti,ab.
245. (Sao Tome adj2 Principe).ti,ab.
246. Senegal.ti,ab.
247. Sudan.ti,ab.
248. (Zambia or Northern Rhodesia).ti,ab.
249. (Zimbabwe or Southern Rhodesia).ti,ab.
250. (Botswana or Bechuanaland or Kalahari).ti,ab.
251. (Equatorial Guinea or Spanish Guinea).ti,ab.
252. (Gabon or Gabonese Republic).ti,ab.
253. (Mauritius or Agalega Islands).ti,ab.
254. Namibia.ti,ab.
255. (Benin or Dahomey).ti,ab.
256. (Burkina Faso or Burkina Fasso or Upper Volta).ti,ab.
257. (Burundi or Ruanda-Urundi).ti,ab.
258. (Central African Republic or Ubangi-Shari).ti,ab.
259. Chad.ti,ab.
260. (((Democratic Republic or DR) adj2 Congo) or Congo-Kinshasa or Belgian Congo or Zaire or Congo Free State).ti,ab.
261. Eritrea.ti,ab.
262. (Ethiopia or Abyssinia).ti,ab.
263. Gambia.ti,ab.
264. (Guinea not (New Guinea or Guinea Pig* or Guinea Fowl or Guinea-Bissau or Portuguese Guinea or Equatorial Guinea)).ti,ab.
265. (Guinea-Bissau or Portuguese Guinea).ti,ab.
266. Liberia.ti,ab.
267. (Madagascar or Malagasy Republic).ti,ab.
268. (Malawi or Nyasaland).ti,ab.
269. Mali.ti,ab.
270. (Mozambique or Mocambique).ti,ab.
271. (Niger not (Aspergillus or Peptococcus or Schizothorax or Cruciferae or Gobius or Lasius or Agelastes or Melanosuchus or radish or Parastromateus or Orius or Apergillus or Parastromateus or Stomoxys)).ti,ab.
272. (Rwanda or Ruanda).ti,ab.
273. (Sierra Leone or Salone).ti,ab.
274. (Somalia or Somaliland).ti,ab.
275. South Sudan.ti,ab.
276. (Tanzania or Tanganyika or Zanzibar).ti,ab.
277. (Togo or Togolese Republic or Togoland).ti,ab.
278. Uganda.ti,ab.
279. Seychelles/
280. Seychelles.ti,ab.
281. Sahel.ti,ab.
282. Reunion/
283. (reunion adj1 island).ti,ab.
284. or/223-283
285. 139 and 172 and 222 and 284
286. limit 285 to yr=“2010 -Current”
287. remove duplicates from 286

## Footnotes

1 The Blueprint list includes Disease X - a hypothetical pathogen currently unknown to cause human disease.

